# Ultrasound-Guided Needling and Lidocaine Plus Sterile Water Injection in Lumbar Spinal Stenosis

**DOI:** 10.1101/2021.02.03.21251072

**Authors:** Carl PC Chen, Areerat Suputtitada, Krit Pongpirul

**Affiliations:** Department of Physical Medicine & Rehabilitation, Chang Gung University, Guishan District, Taoyuan City, Taiwan; Department of Rehabilitation Medicine, Faculty of Medicine, Chulalongkorn University and King Chulalongkorn Memorial Hospital, Bangkok, Thailand; Department of Preventive and Social Medicine, Faculty of Medicine, Chulalongkorn University, Bangkok, Thailand; Department of Health, Behavior and Society, Johns Hopkins Bloomberg School of Public Health, Baltimore, MD, USA

**Keywords:** spinal injection, low dose lidocaine, ultrasound-guided, spinal stenosis, desensitization, one needle insertion

## Abstract

**Objective:** To study the effect of ultrasound-guided (USG) needling and lidocaine plus sterile water injections in lumbar spinal stenosis (LSS).

**Method:** This retrospective study examined data of LSS patients who received USG lidocaine injection to lumbar facets, medial branches to facet joints, and multifidus muscles with one needle insertion at 4 spinal levels.

**Results:** A total of 213 lumbar spinal stenosis patients—104 patients received USG needling and 1% lidocaine without adrenaline 2 ml plus sterile water 10 ml injection (Group A) and 109 patients received 1% lidocaine without adrenaline 6 ml injection (Group B)—for 4 times, once per week. The VAS of chronic low back pain, radicular pain, claudication, and walking ability of both groups at 3, 6, and 12 months were significantly better than the baseline. Group A reported significantly better chronic low back pain, radicular pain, claudication, and walking ability at 3, 6, and 12 months than Group B.

**Conclusions:** USG needling and 1% lidocaine without adrenaline 2 ml plus sterile water 10 ml injection to lumbar facets, medial branches to facet joints, and multifidus muscles with one needle insertion at 4 spinal levels, once a week for 4 weeks can improve low back pain, radicular pain and gait ability in LSS with long term pain relief at least 12 months.

**Highlights:**

- There is evidence of the long-lasting effectiveness of local anesthetic alone for chronic spinal pain on noxious peripheral stimulation, phenotypic changes for neuronal plasticity, and neurotransmitter release responsible for secondary hyperalgesia.
- USG needling and amount of safety solution as 1% lidocaine without adrenaline 2 ml plus sterile water 10 ml is effective for treating chronic low back pain, radicular pain, and claudication in LSS at least 12 months.
- These clinical outcomes should be the effects of peripheral and central desensitization. The other possible effect is the mechanical removal of fibrosis and calcification at lumbar facets, medial branches to facet joints, and multifidus muscles.

## Introduction

Lumbar spinal stenosis (LSS) is a spinal degenerative condition with an increasing prevalence among older ages. Lumbar surgery for LSS has been the most utilized treatment worldwide. The surgical procedures lead to high expenses, mild to severe complications, and repeated surgery with long-term failback syndrome ^1^. LSS patients experience rapid low back pain, numbness, and claudication reduction after surgery. Within 60 months, they always experience low back pain and difficulty in walking ^2^. A systematic review comparing the surgical and nonsurgical treatments is still inconclusive ^3^. LSS may result from disc herniation, degenerative spine disorders, compression deformities, congenital spine disorders, arthritic conditions, tumors, trauma, conditions leading to herniated discs, calcification deposits, and thickened ligaments. Magnetic Resonance Imaging (MRI) evidence of LSS has no good correlation with the symptoms ^4^. The standard of care for spinal stenosis has still been inconclusive. Current evidence reveals small support in epidural steroid injection (ESI) for long-term relief of neurogenic claudication ^4^.

Recently, evidence of using ultrasound-guided (USG) injection is increasing. USG injection has the benefit of outpatient setting, less invasive, less complication, less expense, but importantly need training and experiences of physicians ^4-10^. For long-term effectiveness of the lumbar spine, there is Level II evidence for radiofrequency neurotomy and lumbar facet joint nerve blocks whereas the evidence for lumbosacral intraarticular injections is Level III. For long-term effectiveness of the cervical spine, there is Level II evidence for cervical radiofrequency neurotomy and cervical facet joint nerve blocks, and the evidence for cervical intraarticular injections is level IV. For long-term effectiveness of the thoracic spine, there is Level II evidence for thoracic facet joint nerve blocks and the evidence for radiofrequency neurotomy is Level IV ^9^.

We hypothesize that spinal stenosis can be effectively treated if we can mechanically remove the fibrosis and calcification using only USG needling and further washout by injecting an amount of lidocaine plus sterile water. This innovative treatment could be used in clinical practice but requires high physician diagnostic and interventional USG skills. We published the innovative technique of USG injections at the facet joint, medial branch to the facet joint, and multifidus muscle in one needle ^5^.

## Material and methods

We conducted a retrospective analysis from the medical records of all LSS patients who underwent USG spinal injection from one injector who is the experienced USG intervention, from 1^st^ January 2019 to 31^st^ May 2020.

The inclusion criteria were as the followings: 1) patients with age is beyond 60 years old; 2) patients with characteristic symptoms of chronic low back pain and LSS as neurogenic claudication; 3) patients with MRI findings of LSS; 4) patients who underwent USG lidocaine injections to lumbar facets, medial branches to facet joints and multifidus muscles at bilateral L3-4, L4-5 levels; The exclusion criteria were as the followings: 1) patients treated with pain killer, rehabilitation, other lumbar interventions, back surgery; 2) patients without complete medical records.

All patients received USG injections at the facet joint, medial branch to facet joint, and multifidus muscle in the one needle ^5^, totally 4 needles each session, 2 needles on each side at L3-4 and L4-5 levels, once a week, every week for 4 weeks. During the 4-week treatment period, the patients were asked to avoid inflammation from injury at lumbosacral facet joints and related areas by avoiding objects lifting, back and abdominal exercises, prolong sitting, prolong walking and standing more than 30 minutes, avoid message and any therapy for back pain. The patients were encouraged to do only walking exercises 20 minutes per day. We allocated the patients who received needling and 1% lidocaine without adrenaline 2 ml plus sterile water 10 ml injection, 1 ml at the medial branch, 1 ml at facet joint and 1 ml at multifidus muscles, 3 ml each level as the Group A and the patients who received 1% lidocaine without adrenaline 6 ml injection, 0.5 ml each site, 1.5 ml each level as Group B. The core muscles stretching and strengthening exercises were prescribed at 2 months till 12 months in both groups.

Self-reported low back pain, radicular pain, claudication, and gait ability were obtained at 6 timepoints: pre-injection (baseline), immediately after the injection, one-week, 3-month, 6-month, and 12-month follow up. The self-report pain was measured by the Visual Analogue Scale (VAS) whereas gait ability was measured by the walking distance before calf pain.

### Statistical Analysis

All data were statistically analyzed using the Statistical Program for Social Sciences (SPSS) version 17 (SPSS Inc, Chicago). Descriptive statistics were displayed with mean, range, and standard deviation. Normality tests were done by Kolmogorov-Smirnov test. Intergroup comparisons were done with Mann Whitney U test, and the comparison within each group was tested using Wilcoxon signed-rank test.

### Ethical Consideration

This study was approved by Chang Gung Medical Foundation Institutional Review Board (IRB number 201901477A3). The written consent forms were exempted from the IRB.

## Results

A total of 316 LSS patients were assessed for eligibility. The Strengthening the Reporting of Observational Studies in Epidemiology (STROBE) flowchart is shown in Figure 1. One-third of the patients were excluded. Of the remaining 213 patients, 104 patients were in Group A and 109 patients were in group B. The mean age and gender in Group A and B were 69.78±9.25 years, range 61-90 years, 56.73% females and 68.21±8.57 years, range 61-91 years, 52.29% females, respectively (Table 1).

**Table 1:** Baseline Characteristics of 213 patients.

| Total Number of Patients | Group A<br>(104) | Group B<br>(109) |
| --- | --- | --- |
| Female (%) | 59 (56.73%) | 57 (52.29%) |
| Age (Mean± SD) (years) | 69.78±9.25 | 68.21±8.57 |
| VAS of back pain<br>(Mean± SD) | 8.22±1.56 | 8.73±1.45 |
| VAS of radicular pain<br>(Mean± SD) | 8.56±1.12 | 8.44±1.28 |
| VAS of neurogenic claudication<br>(Mean± SD) | 8.78±0.98 | 8.56±0.43 |
| Walking distance (meters) | 253±24 | 267±41 |
| Radicular pain |  |  |
| - Right | 33 (33%) | 35 (35%) |
| - Left | 49 (49%) | 42 (42%) |
| - Both | 18 (18%) | 23 (23%) |

**Figure 1.**
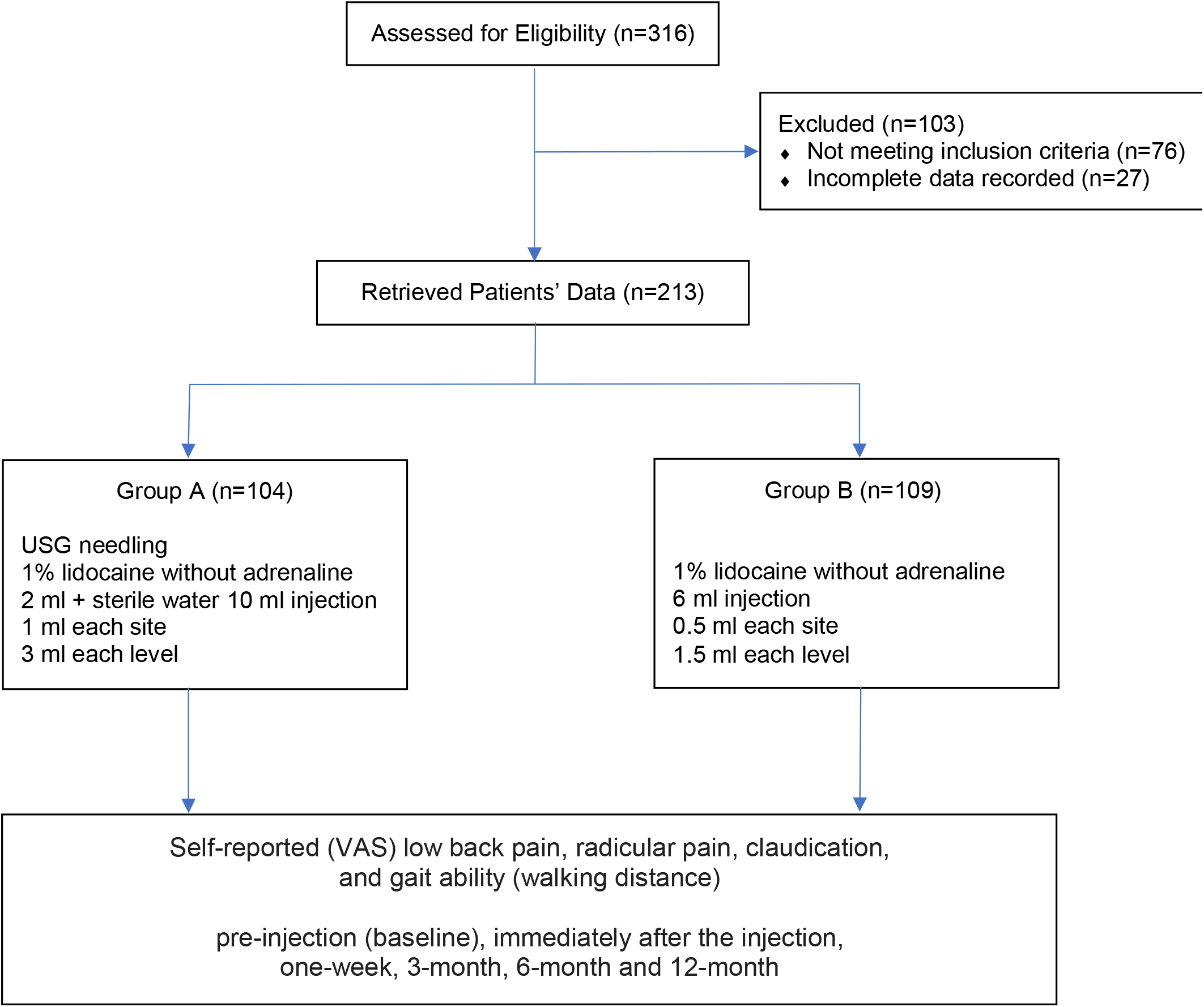
STROBE Flow Chart.

The outcome measurements were presented in Table 2. The VAS of chronic low back pain, radicular pain, claudication, and walking ability of both Groups at 3 months, 6 months, and 12 months were significantly better than the baseline. The VAS of chronic low back pain, radicular pain, claudication, and walking ability of Group A at 3 months, 6 months, and 12 months were significantly better than Group B. No serious effect was found.

**Table 2.**
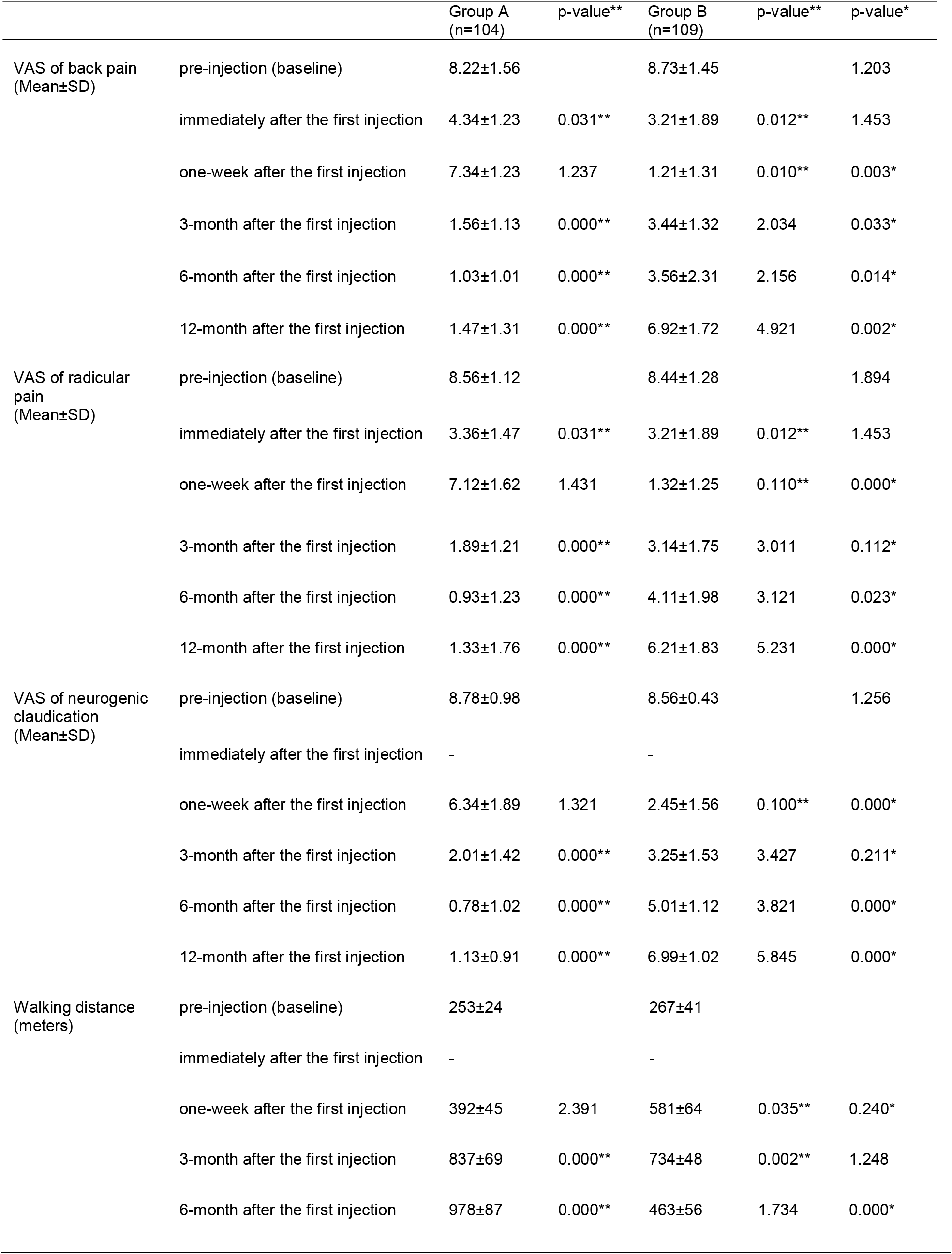
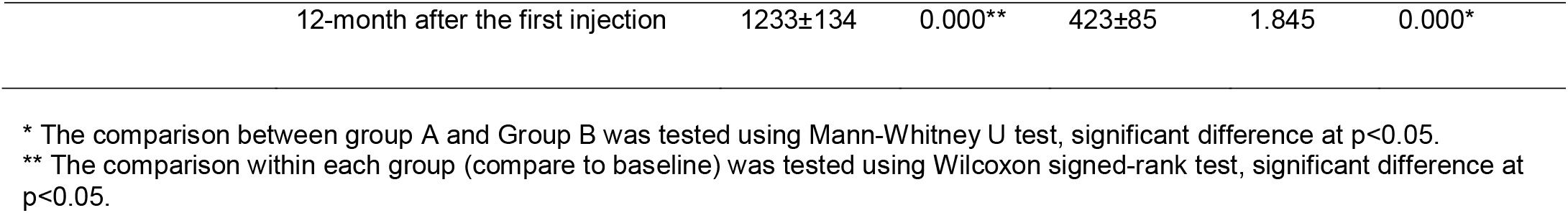
Result of Treatments.

## Discussion

LSS is the condition of spinal canal narrowing, leading to chronic low back pain, radicular pain to the hip, buttock, leg, calf pain while walking, and lower extremity weakness called neurogenic claudication. The leg pain, numbness, and weakness are associated with the lumbar nerve roots entrapment. The injection with local anesthetics and steroids into facet joints, under fluoroscopy and computed tomography (CT) guidance, for LBP by facet joint sprain or degenerative changes is established for facet joint syndrome. The evidence of using USG injection are increasing since the benefit of outpatient setting, less invasive, less complication, less expense, but importantly need training and experiences of physicians ^4-10^. The pathophysiology of facet joint syndrome is a chronic inflammatory process of facet joints and fibrosis with calcification developed afterward. The facet joint syndrome together with degenerative disc or herniated disc are progressing to be spinal stenosis ^1-4^.

The evidence showed that blocking the medial branch of facet joint with local anesthetic alone lead to suppression of nociceptive discharge, blocking of the sympathetic reflex arc and the axonal transport, peripheral and central desensitization, phenotypic changes for neuronal plasticity, and neurotransmitter release responsible for secondary hyperalgesia for a long period of time. and also anti-inflammatory effects ^4; 6-11^ We published the technique of USG injections at the medial branch to the facet joint ^12^.^12^ There are evidences showing local anesthetic effects similar to steroids ^4; 8-11^. There are evidences of the effectiveness of facet joint injection in lumbar spinal stenosis ^13-16^. It has fewer side effects than intraspinal injections due to its direct access to facet joints through paraspinal muscles.

The systematic review ^9^ reported sodium chloride solution injected into an intraarticular space has similar effects as a local anesthetic with a steroid. Moreover, the intraarticular steroid is not an effective therapy. There are different effects from different solutions of injections such as local anesthetic, normal saline, dextrose; and whenever any solution is injected into the disc, facet joint, or multifidus muscle. There are evidences of a small volume of local anesthetic or normal saline abolishing muscle twitch induced by a low current (0.5 mA) during electrode location. Moreover, there is spinal cord involvement via placebo analgesia. Epidural sodium chloride injection causes a significant improvement in pain and function. Besides the accuracy of diagnostic facet joint nerve blocks include local anesthetic effect, there are non-specific effects resulting in positive results of 2 years ^9^.

There are evidences of the desensitization in dilution of local anesthetic with water for safety concern in high-risk patients for myofascial pain syndrome. There was no side effect of sterile water plus local anesthetic agent injecting in muscles for peripheral desensitization in the myofascial pain syndrome. We also injected at trigger points of multifidus muscles, which caused desensitization as well ^17; 18^.

This innovative therapy requires the precision of needling at facet joint since we are not using steroids or any drug for decreasing pain and inflammation. Our target goal is removing the fibrosis and calcification around the facet joint which entraps nerve roots, blood vessels causing radicular pain, claudication, and even leg cramp. This innovative therapy provides gliding of facet joint and increasing vascular supply after all calcification and fibrosis are washed out from facet joints. We use low-dose lidocaine since we also want an anesthetic effect and also block the medial branches. Besides the mechanical removal by needling and wash out by water jet effect of low dose lidocaine plus sterile water which increasing the facet joints motion, there are the effects of peripheral and central desensitization for chronic low back pain and radicular pain relief. The walking ability was increasing because of improvement of spinal nerve function and pain relief.

### Study limitation

This was a retrospective review of data from one institution and one experienced injector resulting in a potentially biased sample, which limited generalizability. A prospective multi-center randomized controlled trial should be done to confirm the effectiveness of this innovative USG intervention. The ultrasonography should be used as the objective outcome for fibrosis and calcification at baseline, during and after complete treatment, and follow up period.

## Conclusions

USG needling and further injection with 1% lidocaine without adrenaline plus sterile water to wash out the fibrosis and calcification around the lumbar facet joints, medial branches to facet joints, and multifidus muscles is effective for at least 12 months.

## Data Availability

Data is available upon reasonable request.

## Acknowledgments

Not Applicable

## Disclosure

The authors report no conflicts of interest in this work.

## Notes

### Competing Interest Statement

The authors have declared no competing interest.

### Funding Statement

This study received no funding support.

